# A Short Plus Long-Amplicon Based Sequencing Approach Improves Genomic Coverage and Variant Detection In the SARS-CoV-2 Genome

**DOI:** 10.1101/2021.06.16.21259029

**Authors:** Carlos Arana, Chaoying Liang, Matthew Brock, Bo Zhang, Jinchun Zhou, Li Chen, Brandi Cantarel, Jeffrey SoRelle, Lora V. Hooper, Prithvi Raj

## Abstract

High viral transmission in the COVID-19 pandemic has enabled SARS-CoV-2 to acquire new mutations that impact genome sequencing methods. The ARTIC.v3 primer pool that amplifies short amplicons in a multiplex-PCR reaction is one of the most widely used methods for sequencing the SARS-CoV-2 genome. We observed that some genomic intervals are poorly captured with ARTIC primers. To improve the genomic coverage and variant detection across these intervals, we designed long amplicon primers and evaluated the performance of a short (ARTIC) plus long amplicon (MRL) sequencing approach. Sequencing assays were optimized on VR-1986D-ATCC RNA followed by sequencing of nasopharyngeal swab specimens from five COVID-19 positive patients. ARTIC data covered >90% of the virus genome fraction in the positive control and four of the five patient samples. Variant analysis in the ARTIC data detected 67 mutations, including 66 single nucleotide variants (SNVs) and one deletion in ORF10. Of 66 SNVs, five were present in the spike gene, including nt22093 (M177I), nt23042 (S494P), nt23403 (D614G), nt23604 (P681H), and nt23709 (T716I). The D614G mutation is a common variant that has been shown to alter the fitness of SARS-CoV-2. Two spike protein mutations, P681H and T716I, which are represented in the B.1.1.7 lineage of SARS-CoV-2, were also detected in one patient. Long-amplicon data detected 58 variants, of which 70% were concordant with ARTIC data. Combined analysis of ARTIC +MRL data revealed 22 mutations that were either ambiguous (17) or not called at all (5) in ARTIC data due to poor sequencing coverage. For example, a common mutation in the ORF3a gene at nt25907 (G172V) was missed by the ARTIC assay. Hybrid data analysis improved sequencing coverage overall and identified 59 high confidence mutations for phylogenetic analysis. Thus, we show that while the short amplicon (ARTIC) assay provides good genomic coverage with high throughput, complementation of poorly captured intervals with long amplicon data can significantly improve SARS-CoV-2 genomic coverage and variant detection.

## Introduction

Severe acute respiratory syndrome coronavirus 2 (SARS-CoV-2), the causative pathogen for COVID-19 disease, continues to impact the global population with a growing number of variants (1). Community spread is the predominant mechanism leading to the increasing incidence of COVID-19 disease world-wide. SARS-CoV-2 genome sequencing data suggest that several novel variants and regional strains are emerging in the United States (2). Whole genome sequencing (WGS) analysis of these viruses enables high resolution genotyping of circulating viruses to identify regionally emerging strains (3, 4). Genome sequencing is a powerful tool that can be used to understand the transmission dynamics of outbreaks and the evolution of the virus overtime (5). Phylogenetic analysis of WGS data can reveal a virus’s origin and genetic diversity of circulating strains of the virus (6, 7). With the ongoing pandemic, SARS-CoV-2 is getting ample amounts of opportunities to replicate and incorporate new mutations that can potentially impact virus characteristics such as transmissibility as reported in the cases of the B.1.1.7 lineage in England and the B.1.351 lineage in South Africa (8-10). In addition, the abundance of mutations in new strains can also impact the performance of diagnostic and research methods that were developed based on the original reference genome from the beginning of the pandemic last year (11, 12). Therefore, methods and strategies of virus detection and genome sequencing need to be updated.

The ARTIC network protocol is one of the most widely used methods to sequence the SARS-CoV-2 genome (7, 13). Two pools of primers in this assay amplify multiple short amplicons to assemble the entire genome. An increasing number of mutations in emerging strains poses one potential challenge to this strategy, as mutated primer binding sites may cause amplicon dropout or uneven sequencing coverage resulting in loss of information or inaccurate data. To address this issue, we developed a new approach that amplifies each specimen using both, short and long amplicon primers to uniformly capture the entire viral genome. This approach offers two potential benefits. First, a smaller number of primers needed to amplify the entire genome reduces the chance of encountering a mutated site. Second, besides single nucleotide changes, deletions and insertions can be captured more effectively with longer amplicons and sequencing reads. Here, we present our approach to supplement ARTIC’s short amplicon sequences with long-amplicons to generate more complete and high-quality sequencing data for mutation detection and phylogenetic analysis (Figure 1A).

**Figure 1.**
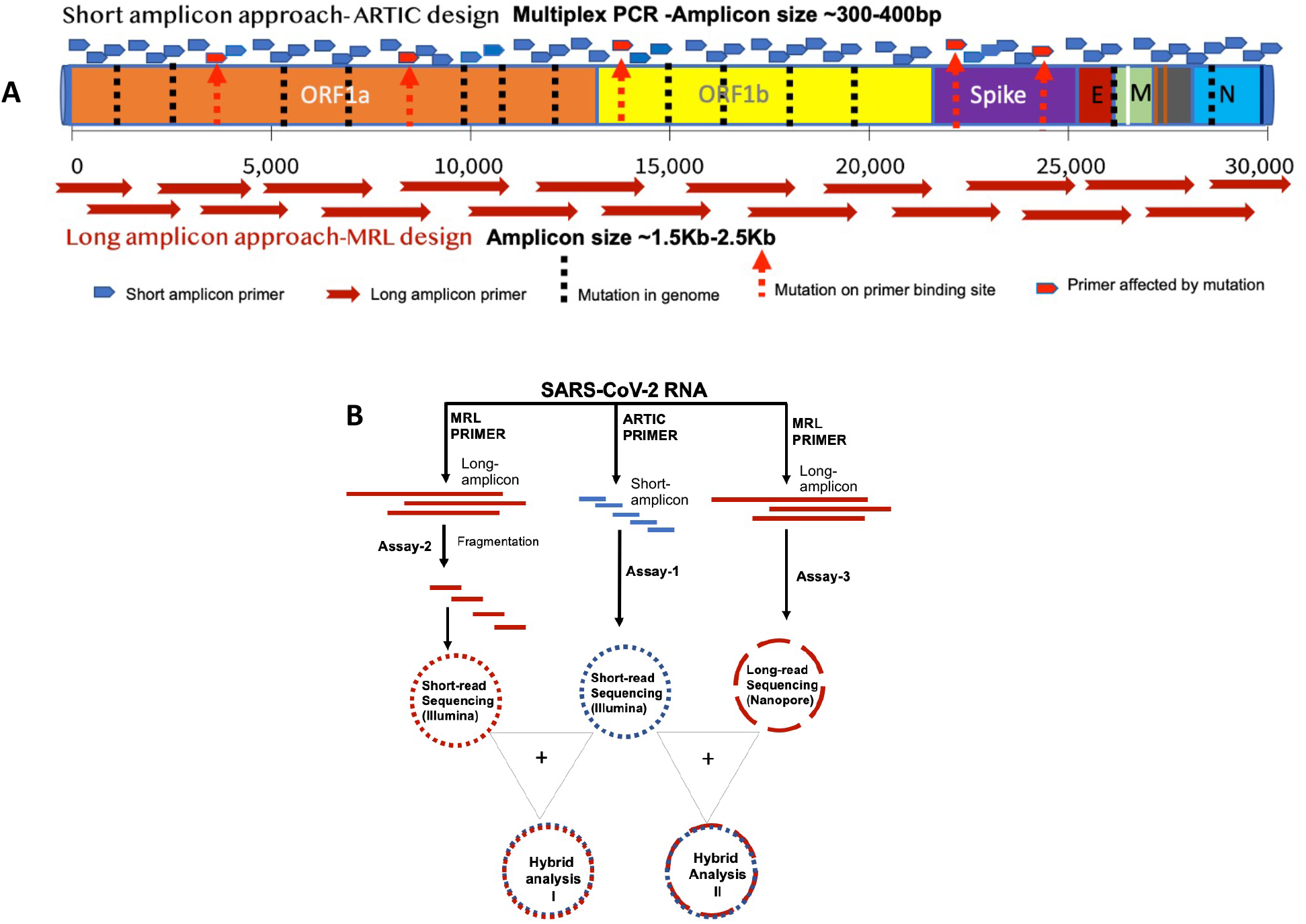
Study rationale and assay design. Panel A illustrates the study’s rationale and sketches the layout of the ARTIC and MRL primer pools across the SARS-CoV-2 genome. Small and long arrows indicate short and long amplicons, respectively. Red arrows indicate a mutation with potential to alter the primer binding site. Panel B shows the design of the three assays developed and assessed in the present study. Assay-1 is based on short-amplicons generated with the ARTIC primer pool. Assay-2 is based on long-amplicons made with MRL primers followed by short-read sequencing on MiSeqDx. Assay-3 is based on long-amplicons made with MRL primers followed by long read sequencing on MinION platform.

## Materials and Methods

### a. Sample

We used ATCC VR-1986D as our positive control RNA and a human dendritic cell RNA as our negative control to test our own primer design (MRL-Primer) and ARTIC primers. Next, we tested RNA from five SARS-CoV-2 virus positive nasopharyngeal swabs in universal transport media. Samples were de-identified and analyzed with a waiver from UT Southwestern Institutional Review Board.

### b. Primers

We designed our own set of primers that amplify long-amplicons spanning 1.5-2.5Kb of the SARS-CoV-2 viral genome. Details on our primer sequences are provided in Table 1. ARTIC data was generated using ARTIC nCoV-2019 V3 Panel primers (13) purchased from Integrated DNA technologies (IDT).

### c. Assays and protocol

We developed three assays to sequence the SARS-CoV-2 genome and assessed their performance in the present study (Figure 1B). In Assay-1, virus genome was amplified using the ARTIC primer pool only and sequenced on the MiSeqDx Illumina platform. The second assay is used only on our own primer design (MRL Primer) that generates 19 long amplicons of 1.5-2.5Kb in size to capture the complete genome of the virus. These long amplicons were then fragmented into 300-500bp sizes and sequenced on the MiSeqDx platform. The third assay also used only on our own primer design (MRL Primer), but the long-amplicons were directly sequenced on the long-read sequencing platform, MinION, from Oxford Nanopore Technology (ONT). Finally, the performance of the individual assays as well as the combined assays, (Hybrid I & Hybrid II), were assessed.

### Step 1: RNA extraction and quality control

RNA was extracted from nasopharyngeal swabs using the Chemagic Viral DNA/RNA 300 Kit H96 (Cat# CMG-1033-S) on a Chemagic 360 instrument (PerkinElmer, Inc.) following the manufacturer’s protocol. A sample plate, elution plate, and a magnetic beads plate were prepared using an automated liquid handling instrument (Janus G3 Reformatter workstation, PerkinElmer Inc). In brief, an aliquot of 300µl from each sample, 4µL Poly(A) RNA, 10µL proteinase K and 300µL lysis buffer 1 were added to respective wells of a 96 well plate. The sample plate, elution plate, (60µL elution buffer per well) and magnetic beads plate (150µL beads per well) were then placed on a Chemagic 360 instrument and RNA was extracted automatically with an elution volume of 60µL. The final quality and quantity of RNA was estimated using an Agilent 2100 Bioanalyzer System and Agilent RNA 6000 Nano Kit, catalog number 5067-1511; or Agilent RNA 6000 Pico kit, catalog number 5067-1513. A Quant-it Picogreen dsDNA assay kit and PerkinElmer Victor X3 2030 multilabel reader were used to measure cDNA concentration.

### Step 2: cDNA synthesis and PCR amplification of SARS CoV-2 genome using gene-specific primers

We used Invitrogen SuperScript™ III One-Step RT-PCR System with Platinum™Taq High Fidelity DNA Polymerase (Catalog Number: 12574-035) to make cDNA. SARS Cov-2 gene-specific primer set *MRL*-design and ARTIC design were synthesized from IDT. The MRL primer set included two primer pools (19 pairs, about 1.5Kb to 2.5Kb/ amplicon, Tm 59-60°C), whereas ARTIC CoV-2019 primer pools included 109 pairs, about 400nt/ amplicon, Tm 60-62°C. Both primers were used to amplify ATCC VR-1986D genomic RNA from severe acute respiratory syndrome-related coronavirus 2 Positive control. We also used a negative control of Human RNA extracted from monocyte-derived dendritic cells (MDDCs) to test our assay. We started with 0.1 ng of ATCC VR-1986D RNA (8000 genome copies), 100 ng of human MDDCs RNA, and 3 ul of Covid -19 patient RNA (RT-PCR Ct value 30) as an input RNA amount per each cDNA synthesis reaction. Reverse transcription was performed at 50°C for 30 min, followed by denaturation at 94°C for 2 min. PCR amplification involved 35 cycles (95°C for 30 s, 55°C for 1 min, 68°C for 4.5 min) followed by a final extension at 68°C for 10 min. Reaction products from two primer pools were combined and a bead-based cleanup was performed. Agencourt AMPure XP beads by Beckman Coulter (Catalog# A63881) were used for purification. Then, the cDNA quantity was measured using the Picogreen method. Quant-iT™ PicoGreen dsDNA Assay kit by Invitrogen with Catalog # P7589 and a PerkinElmer plate reader (PerkinElmer Victor X3, 2030 Multilabel Reader) were used in the assessment.

### Step 3: NGS workflow-Amplicon Library Preparation and quality control

We used the Kapa HyperPlus Library Preparation Kit (Catalog #KK8514) to construct our sequencing libraries. The cDNA input amount for each library preparation was between 100-500 ng due to the limitation of cDNA quantity. We started with a 15-minute 37°C enzymatic fragmentation for the cDNA amplicons generated by MRL-primer because of the large amplicon size (1.5Kb-2.5Kb). Since ARTIC amplicons sizes were already around 400bp, no fragmentation was performed on these replicates. Then end-repair, A-tailing, and UMI adapter ligation were performed on the amplicons. After the ligation step, we did double-sided size selection with AMPure XP beads followed by 4 to 8 PCR cycles to amplify adapter ligated fragments. Finally, the amplified libraries were purified by AMPure XP beads to form the final libraries. The libraries’ quantity was measured with Picogreen, and the quality was verified with a Bioanalyzer (Agilent DNA 1000 kit, catalog # 5067-1504 or Agilent High Sensitivity DNA kit, Catalog # 5067-4626). In addition, we did qPCR to check the adapter ligation efficiency using Applied Biosystems 7500 Real-Time PCR instrument.

### Step 4: Sequencing

About 10pM barcoded libraries were sequenced on MiSeqDx sequencer using 500 cycle v2 flow cell kit. About 5% PhiX DNA was added to the sequencing run to increase diversity. The summary of sequencing metrics for all three assays is given in Table 2. After trimming and removing the adapter, quality pass sequencing reads were mapped to the SARS-CoV-2 genome. To facilitate reproducibility, analysis samples were processed using the publicly available nf-core/viralrecon pipeline version 1.1.0 implemented in Nextflow 20.01.0 using Singularity 3.3.0 (10.5281/zenodo.3901628). Briefly, reads were trimmed using fastp, and de novo assembly was performed by spades, metaspades, unicycler, and minia. Variant calling was done by varscan2, ivar, and bcftools using viral genome NC_045512.2 as reference for comparison. Contigs and scaffold metrics were compared using quast and icarus.

### Step 5: Nanopore Library Preparations and Sequencing

The leftover cDNA amplicons from step 2 were used to generate libraries for long-read sequencing analysis on MinION. EXP-NBD104 and SQK-LSK109 kits were used with Oxford Nanopore’s (ONT) native barcoding protocol (Version: NBE_9065_v109_revV_14Aug2019) to construct the libraries. Library preparation was performed according to the manufacturer’s recommended protocol. Native barcoded libraries were pooled in equimolar concentrations and loaded onto a MinION sequencer using a R9.4 flow cell. Sequencing was run for 48 hours. The FAST5 data was basecalled using ONT’s Guppy pipeline.

### Data analysis

FASTQ files from the Illumina MiSeqDx run were generated using bcl2fastq2. Sequencing reads were trimmed using Cutadapt (min Q30, adapter presence, shorter than 50 bases). The presence of host reads were detected using Kraken 2, the host genome version used was GRCh38. Variant calling was done using three methods: varscan2, ivar, and bcftools, and variants were filtered where the BAQ was less than 20 or had a depth less than 10 reads. Minimum allele frequency for calling variants was set to 0.25, max was 0.75. De novo assembly was performed using spades, CoronaSPAdes, metaspades, unicycler and minia. Quast was used to summarize assembly statistics where the reference viral genome was NC_045512.2. Fast5 files from Mk1C runs were base called and demultiplexed with guppy (GPU enabled), PycoQC, FastQC and NanoPlot were used for sequencing QC visualizations. The ARTIC network bioinformatics pipeline (https://github.com/artic-network/artic-ncov2019) was also used to generate a comprehensive view of our Oxford Nanopore long reads. Briefly, reads were mapped to NC_045512.2 with Minimap2 and bams were sorted and indexed. Variants were called using Nanopolish where ploidy was set to 1. The minimum allele frequency was 0.15, the minimum flanking sequence was 10 bases, and at most, 1000000 haplotypes were considered. Combination of Illumina MiSeq paired end reads and Oxford Nanopore Mk1C long reads were assembled using SPAdes, hybrid assembly stats were summarized with Quast. Contiguous scaffolds were visualized with bandage (https://github.com/rrwick/Bandage). Pipeline automation was done by creating Nextflow workflows (v20.01.0).

## Results

### Assay-1: Short-amplicon sequencing results

We generated 1-5 million sequencing reads on each study sample including the positive control ATCC SARS-CoV-2 RNA and five COVID-19 positive patients that had their nasopharyngeal swabs tested by RT-PCR for the presence of SARS-CoV-2. All the patient samples had RT-PCR Ct values <30. The ARTIC primer generated >80% pass filter sequencing reads. The summary of sequencing metrics is given in Table 2. As shown, 99% of sequencing reads on ATCC SARS-CoV-2 RNA mapped to SARS-CoV-2 reference genome (NC_045512.2). Similarly, >90% of sequencing reads in patient samples #NP1-NP4 and about 50% reads in #NP5 mapped to the reference genome (Fig 2A-B). These viral reads in positive control and patient sample NP1-NP4 accounted for >90% of the virus genome, while only 80% of genomic fraction was covered in case of NP5 (Table 2). Variant analysis detected 66 single nucleotide variants (SNVs) and one deletion (Table 3). Of these 66 SNVs, five were present in spike gene region, including nucleotide variants, at position nt22093 (M177I) in sample NP2, position nt23042 (S494P) in sample NP4, position nt23403 (D614G) in NP1-NP5, and position nt23604 (P681H) & nt23709 (T716I) in sample NP4. The mutation at position nt23403 (A → G) is known as the D614G variant which has been shown to alter the fitness of SARS-CoV-2 (14). Interestingly, two of the spike gene variants at nucleotide positions 23604 (P681H) & 23709 (T716I) in sample NP4 represent B.1.1.7 lineage of SARS-CoV-2 (15).

**Figure 2.**
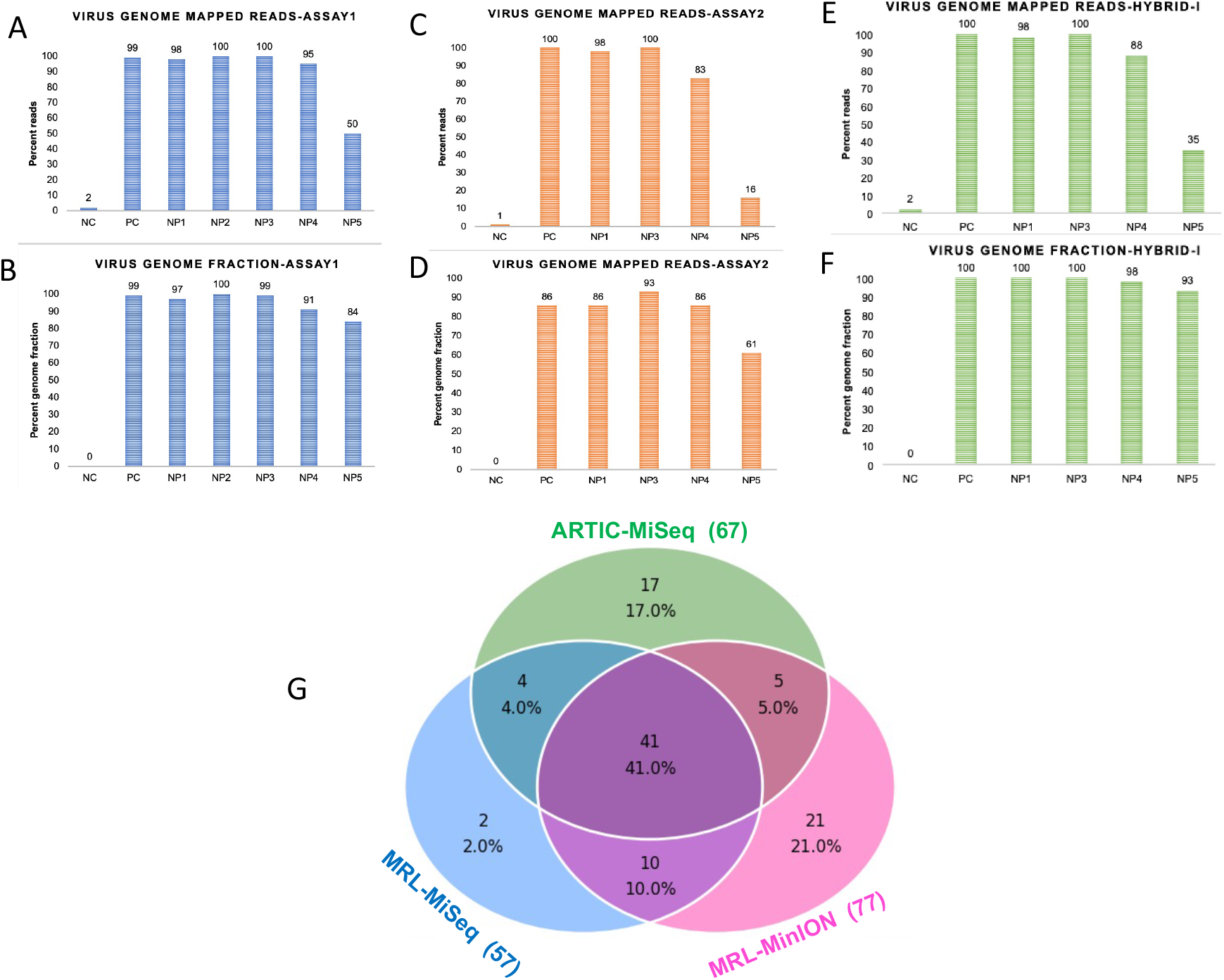
Hybrid analysis based on both the primers improves percentage of virus mapped reads and genomic fraction. Percentage of virus genome mapped sequencing reads (A) and virus genome fraction covered with ARTIC primers (B). NC represents the negative control (Human RNA), PC represents the positive control (VR1986D ATCC SARS-CoV-2 RNA), and NP1-NP5 are the five nasopharyngeal swab RNAs from COVID-19 positive patients. Panels C and D, show the percentage of viral genome mapped sequencing reads (C) and virus genome fraction covered (D) with MRL primers. Data in panels E & F show the percentage of the viral genome mapped sequencing reads (E) and virus genome fraction covered (F) in ARTIC plus MRL or Hybrid data set. The NP2 sample could not be analyzed with MRL primers due to limited sample material. Panel G: This Venn-diagram summarizes the number of mutations that were uniquely or commonly detected by each sequencing assay.

### Assay-2: Long-amplicon based sequencing results

Next, we analyzed the sequencing data generated with MRL primers: the Assay-2. This assay did not include sample NP2 due to insufficient material. The sequencing metrics for this assay are also described in Table 2. Mean sequencing coverage was about 2000X on an average, except for NP5 samples which performed poorly overall. The percentage of viral genome mapped reads and genomic fraction covered with MRL primers was >80% (Figure 2C-D). Variant analysis on assay-2 data identified a total of 57 variants in study samples (Table 4), of which 45 (69%) matched with those called in the short-amplicon data, assay-1. SNP concordance analysis on these 45 variants showed 100% genotype concordance between short and long amplicon data in 4 out of 5 samples. The NP5 sample showed only 91% concordance due to overall poor genomic coverage and data quality. About 9% of the variants in this sample were discordant between assays 1 & 2. Overall, analysis of the data from assay-1 & assay-2 showed that 22 variants were unique to either data set. Further in-depth analysis of the variants that were unique to either assay-1 or assay-2 revealed that the observed discrepancy was largely due to insufficient sequencing read depth to call a variant with analysis pipeline criteria.

### Assay-1 & Assay-2 combined (Hybrid-I) analysis

To improve the sequencing coverage across poorly captured genomic segments in individual assays, we merged the sequencing reads from assay-1 and assay-2 to generate a hybrid assembly and then called the variants again. The hybrid data set has about 7-12 Million reads per sample. As shown, hybrid-I data improved both percentage of virus mapped reads as well as a fraction of covered genome (Figure 2E-F). The number of unique and commonly detected mutations in each assay is summarized in Figure 2G.

Next, analysis of sequencing coverage in short (ARTIC) and long amplicon (MRL) identified genomic intervals that were poorly captured in ARTIC, only assay was performed (Figure 3). As illustrated in Fig.3A, short amplicon data in positive control ATCC RNA and three patient samples (NP1, NP3, NP4) show genomic intervals that have a sudden drop (<10 sequencing reads) in sequencing coverage across the spike and ORF3a gene regions. On the other hand, long-amplicon sequencing data on the exact same set of samples shows more uniform coverage across this region (Fig.3B pink tracks). The red color lines on the coverage tracks indicate the detected mutations in patient samples. As shown, mutations, i.e.,Q57H in the genomic region that have good uniform sequencing coverage, were consistently picked up by both short (Panel 3A) as well as long (Panel 3B) amplicon data. However, a common mutation, G172V, in ORF3a gene at nt25907(G → T), located in the poorly captured region, was missed by the short amplicon data (Panel A), whereas the long-amplicon data did detect this mutation in samples NP1 and NP3 (Panel B). Next, analysis of merged short and long amplicon sequences confirmed this mutation with high confidence in Hybrid data as shown in Fig.3C. Thus, hybrid data improved the detection of 22 mutations that were either ambiguous or not called in ARTIC data due to uneven sequencing coverage (Table 5). Finally, 59 high confidence mutations were identified in our study samples (Table 6).

**Figure 3:**
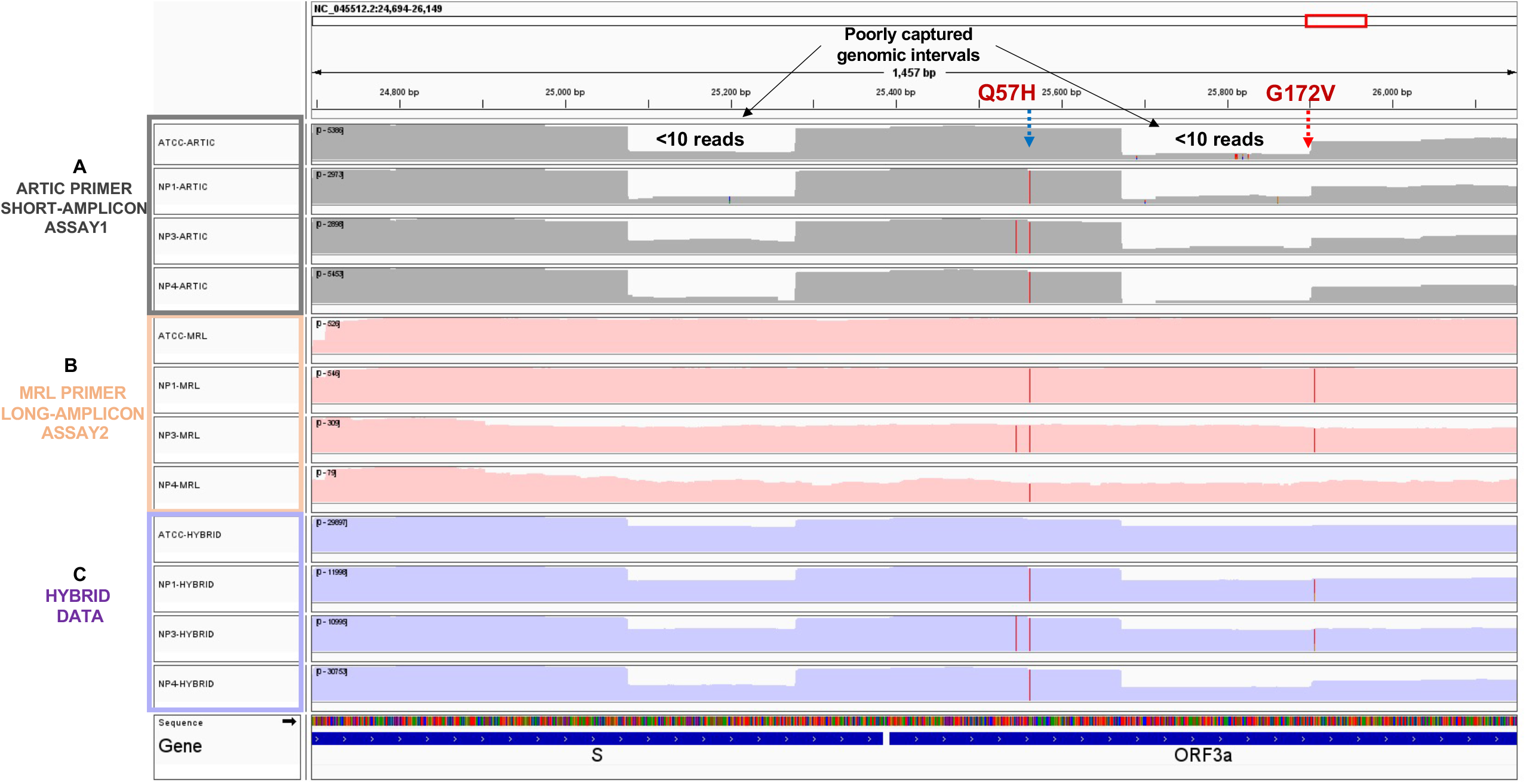
Long-amplicon sequences provide uniform genomic coverage and detects all the mutations. The Integrative genomics viewer (IGV) plot shows sequencing coverage tracks on ARTIC (short-amplicon only-panel A), MRL (long-amplicon only-panel B) and Hybrid (short+long amplicons-panel C). Data is shown on ATCC RNA and three patient samples (NP1, NP3, and NP4). The x-axis shows the genomic position in virus’s genome and the y-axis shows the individual samples. Red colored lines on the sequencing coverage tracks indicate detected mutations. Detection of two mutations Q57H and G172V in ORF3a gene are shown. Black solid arrows point to the poorly captured genomic intervals in ARTIC data set.

### MJ network analysis show interpatient divergence in viral genome

We used the Median-Joining (MJ) Network to illustrate interpatient divergence in viral genome sequences. This analysis was performed on 59 high confidence mutations found in the hybrid data set (Figure 4). Figure 4A shows the distribution of these 59 mutations within the study samples. Although our study sample size is very small to accurately estimate the population frequency of detected mutations, we observed that 11 out of 59 mutations were present in more than 2 specimens, suggesting that they are more common mutations. These common mutations are shown with solid black bars in Figure 4A. Amino acid changes in these common variants are indicated on the top of each bar. The sequence divergence in virus genomes among different samples is illustrated utilizing the median neighbor joining (MJ) algorithm (16) (Fig. 4B). For MJ diagrams, the spheres (termed nodes) represent individual samples in the network. Mutations or single nucleotide polymorphisms (SNPs) that distinguish each node are listed along the line that connects them. These networks are progressive, such that the two nodes at opposite ends of the network are the most divergent (Figure 4B). As shown, ATCC RNA differs from the reference NC_045512.2 by just two mutations (SNP28 and SNP48), whereas all the patient samples differ by multiple mutations that define the network reticulations. As shown, SNP36 (D614G mutation), one of the most common spike gene variants reported in the US population, is present in all of the patient samples (NP1-NP5). Interestingly, sample NP4 carries three additional spike gene mutations (SNP35, SNP37, SNP38). Two of these, P618H and T716I, have been previously reported in the B.1.1.7 lineage spike gene mutations. This analysis allows for the visualization of virus genetic diversity among patients. Next, we performed a lineage prediction analysis using the https://nextstrain.org/ncov/global database. As shown in the MJ network, ATCC RNA is assigned lineage 19B, NP1-20G, NP2-20G, NP3-20G, NP4-19B and NP5-19B.

**Figure 4:**
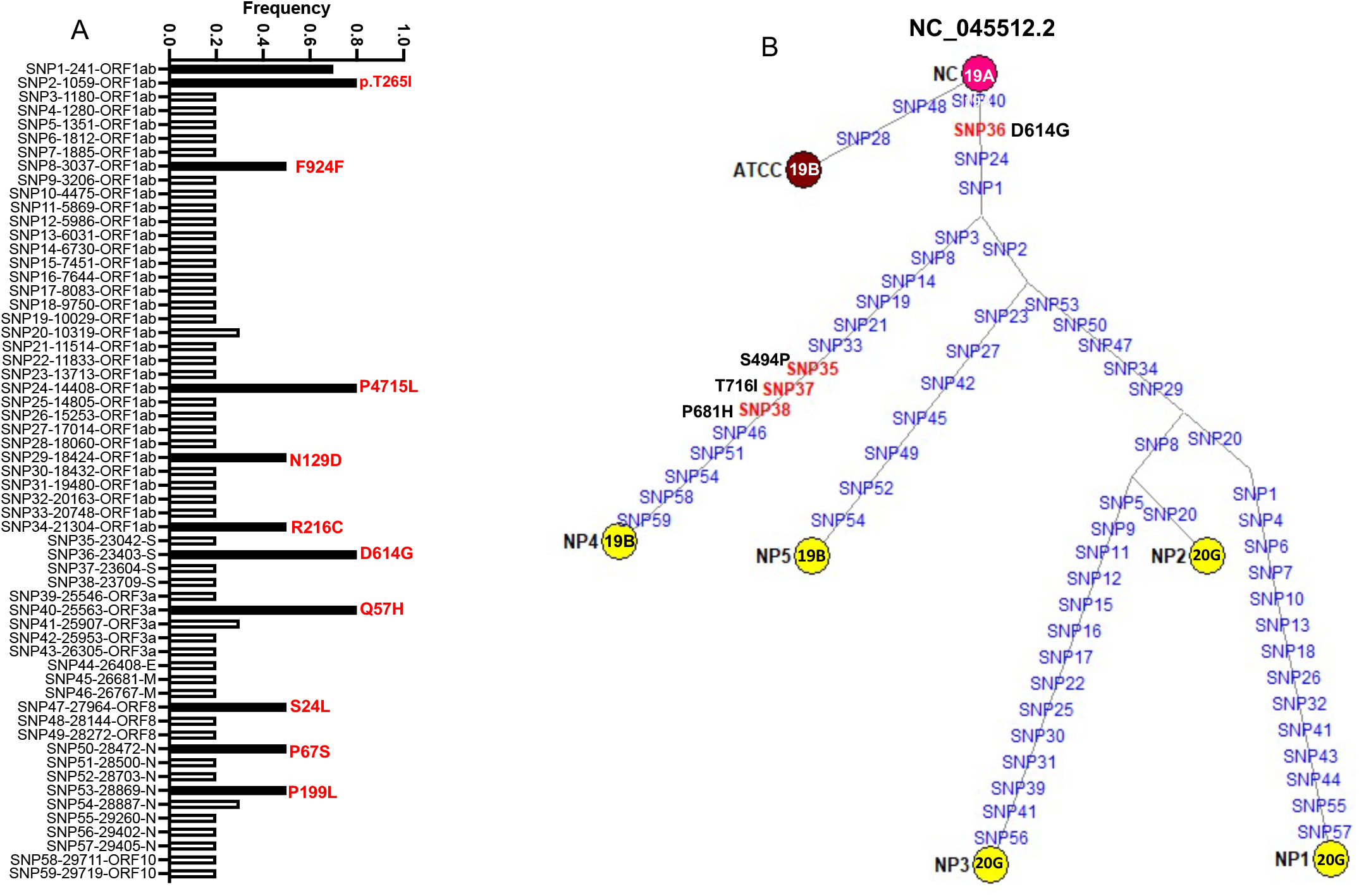
Median-joining (MJ) network analysis on high confidence variants. Panel A shows a frequency plot of 59 high confidence variants within the study samples. The x-axis shows the number of single nucleotide polymorphisms (SNP) or mutations, its position in the genome and the associated gene. The y-axis shows the frequency of a given mutation in the study sample. Each bar represents an individual mutation. In this study’s cohort, 11 high bars show mutations with >50% frequency. Amino acid alterations are shown on the top of each bar for the 10 most common non-synonymous mutations. Panel B shows the Median-joining (MJ) network analysis on 59 mutations plotted in Panel A. Yellow spheres (termed nodes) represent the individual patient samples. The lines connecting the nodes are labeled with the variants or mutations that distinguish viruses in different patients. The length of each network is proportional to the number of variants. Red highlighted variants indicate spike gene mutations. The numbers overlaid on the spheres or nodes indicate the https://nextstrain.org/ncov/global predicted virus lineage for that specimen.

### Assay-3: MRL long-read sequencing analysis on MinION

Given that several deletions have been reported in emerging strains of SARS-CoV-2, it is important that sequencing assays capture deletion-prone genomic intervals appropriately. To assess the performance of our MRL primers with the long-read sequencing workflow, we sequenced the long amplicons from the ATCC positive control and three patient samples (NP1, NP3 & NP4) on MinION R.9.4.1 flow cell for 48 hours. As summarized in Table 2, we generated 500K-1 million sequencing reads on each sample on MinION. More than 80% of sequencing reads were mapped to the viral genome (Figure 5A). The genomic fraction covered was >95% in all four samples (Figure 5B, Table 2). We mapped the nanopore sequencing data to the reference genome and called the variants. In total, 77 mutations were detected in the nanopore sequencing data, of which 46 were concordant with ARTIC assay-1, and 51 were concordant with MRL assay-2 data from Illumina short-read sequencing (Table 7). About 21 mutations were unique to the long-read data from MinION, which needed further validation. Nanopore data detected one insertion in NP1, one deletion in NP3, and one deletion and one insertion in NP4 (Table 7). However, these insertions and deletions were only detected in nanopore data, so further validation and confirmation is needed. We also combined MRL long-read sequences with ARTIC short-read sequences as Hybrid-II data and observed improvement in the percent of reads mapping to the viral genome as well as in overall genomic coverage (Figure 5 C-D). As shown in case of ATCC RNA, we observed that long-read sequencing data has more uniform sequencing coverage across the deletion-prone region of spike gene than short-read ARTIC data. MRL long-read data shows uniform depth of coverage across the 2KB interval in the spike gene region as compared to short-amplicon data for ATCC positive control (Figure 5F). The bottom panel (Fig.5G) shows a snapshot of UCSC genome browser across the spike gene region where several micro and major deletions have been reported recently in emerging lineages of the SARS-CoV-2 virus (Figure 5G).

**Figure 5:**
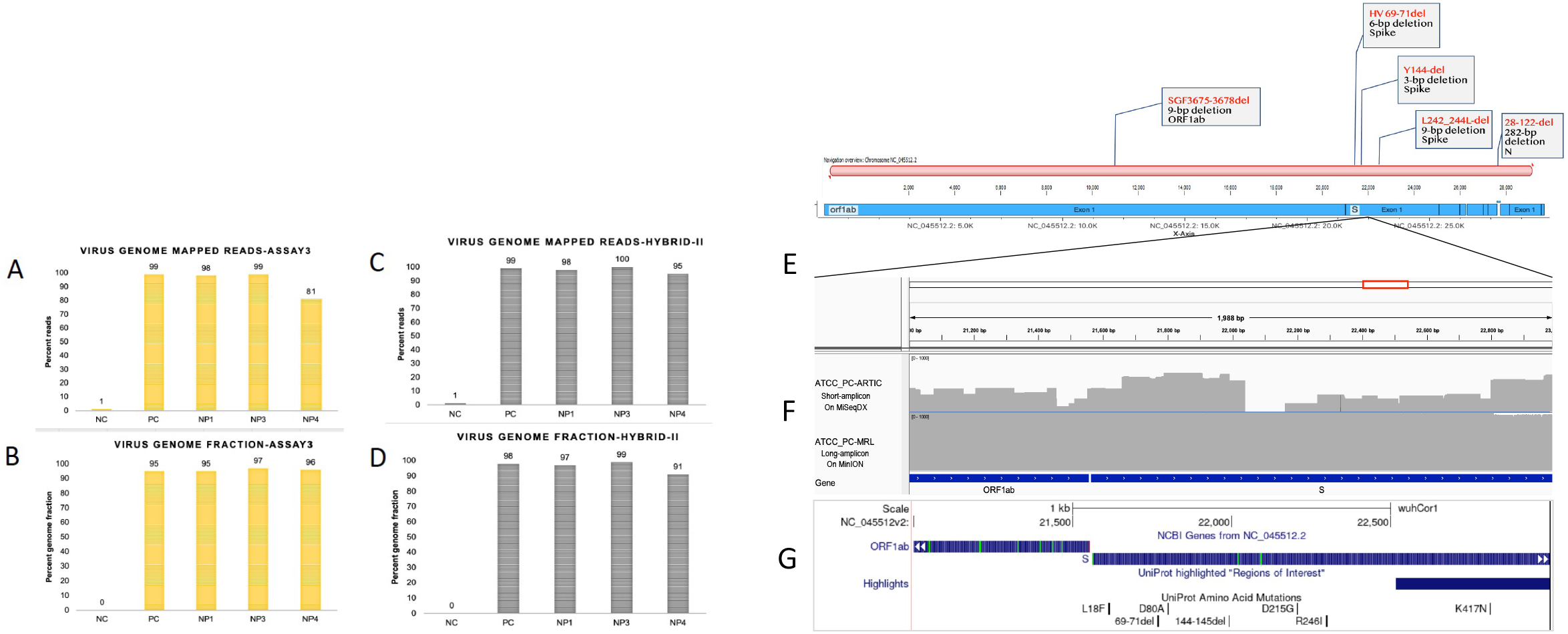
Long-read sequencing provides uniform coverage across deletion-prone region in the virus genome. Panel A shows the percentage of the virus genome mapped sequencing reads (A) and virus genome fraction covered with Nanopore sequencing data (B), in the positive control (VR1986D ATCC SARS-CoV-2 RNA) and NP1, NP3 & NP4 samples. Panels C-D show reads mapped to the virus genome and covered genomic fraction in combined, ARTIC + Long-read (Hybrid-II) data set. Panel E illustrates a gene sketch on known deletions in 2kb region of the spike gene. Panel F shows sequencing coverage across a deletion-prone region of the spike gene in ATCC positive control RNA in short-amplicon ARTIC and long-amplicon MRL data. Top coverage plot on ATCC_PC_ARTIC shows sequencing coverage in short-amplicon on Illumina MiSeqDx platform, whereas second plot show long-read sequencing data on ATCC_PC sample using MRL long-amplicon primers. Panel G shows a UCSC genome browser snapshot across spike gene region to show previously reported deletions in the shown genomic interval of SARS-CoV-2 genome.

## Discussion

High viral transmission in the ongoing COVID-19 pandemic is enabling SARS-CoV-2 to mutate at a faster rate, resulting in an abundance of new mutations in the genome (7, 15, 17, 18). Since most of the currently used PCR primers, protocols, and sequencing strategies were developed on the original reference genome in the beginning of the pandemic, mutations in new strains of the virus might impact the diagnostic and research methods (19). The ARTIC primer pool that amplifies multiple short amplicons in a multiplex-PCR reaction is a widely used method to sequence the SARS-CoV-2 genome (13, 18, 20). However, as no method is perfect, researchers have identified issues with existing short amplicon-based methods and provided potential alternative solutions (13, 21-23). Data from the present study and others show that some key regions of the genome are poorly captured with ARTIC primers (24). This could be due to poor performance of some primer pairs in a large, multiplexed PCR due to either sub-optimal PCR conditions within the assay or emerging mutations on primer binding sites. Our approach to capture and assemble the genome using both short and long-amplicons potentially decreases the chances of missing an interval due to inability of short-amplicon primer to anneal to a mutated site. This improves the genomic coverage, sequencing depth and enables variant calling with high confidence. Accurate detection of all the mutations in the virus genome is important for correct phylogenetic lineage assignment and functional studies.

Our data suggests that sequencing each sample with both short and long amplicon primers can provide more complete and comprehensive genomic coverage to accurately detect true mutations across the genome. The two primer set approach also allows confirmation of low frequency novel mutations and excludes sequencing and analysis related artifacts. We admit that sequencing with two sets of primers would slightly increase the cost of reagents and processing time in the protocol. However, we believe it is worth it for the accuracy and completeness of the genomic data, especially given that several new mutations are emerging in new strains. Although our findings are based on a small number of samples, the presented data provides strong support in favor of our proposed method. Overall, we have demonstrated a new strategy to capture the mutating SARS-CoV-2 genome more accurately and efficiently, allowing for improved analysis of genomic information on evolving strains.

## Data Availability

Sequencing data (FASTQ files) from the present study have been deposited in the NCBI SRA database with accession ID PRJNA729878 for public access.

## Acknowledgments

This study was supported by the UT Southwestern Microbiome Research Laboratory. Authors gratefully acknowledge donors of de-identified patients who provided nasopharyngeal swabs samples for this study. Our sincere thanks to Kaylee D for her diligent proofreading of the manuscript.

## Author contributions

C.A. performed sequencing and bioinformatic analysis, C.L. developed and optimized the sequencing assays on Illumina platform, M.B. and B.Z. developed long-read assays; J.Z. performed sequencing quality control, L.C. extracted RNA from swabs, J.S. provided patient samples for the study, B.C. contributed to data analysis, L.V.H. contributed to manuscript writing and editing, and P.R. conceived and designed the experiments, analyzed data, and wrote the manuscript.

## Ethical approval and patient consent

Review waived by UT Southwestern Institutional Review Board as analyzed specimens were de-identified and were residual material.

---

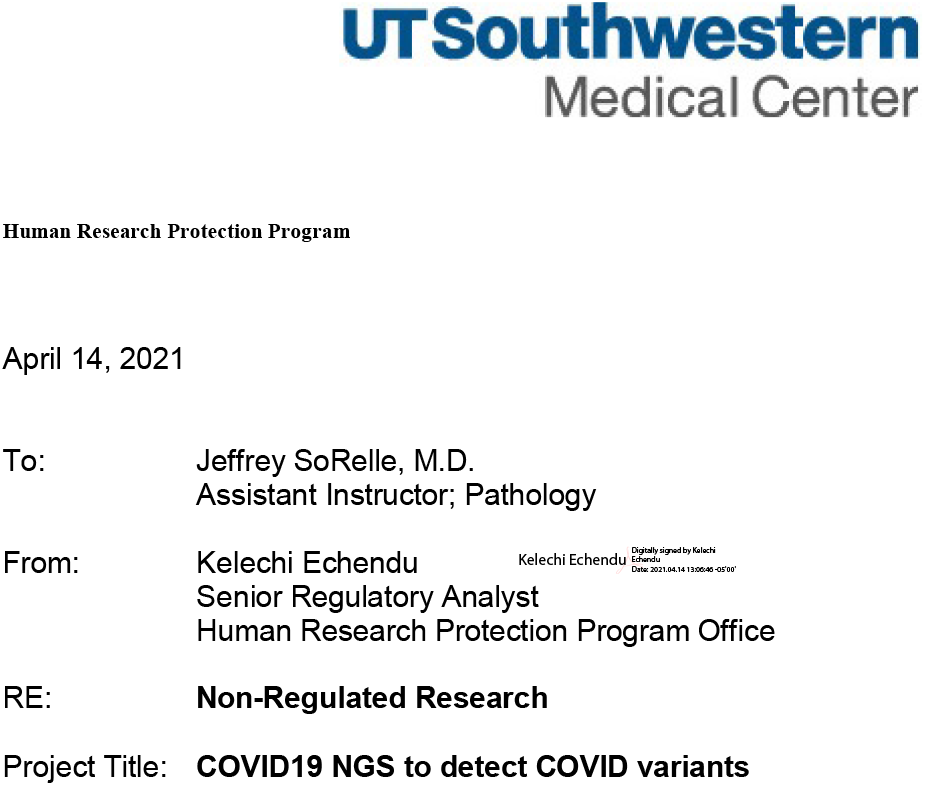

The UT Southwestern Human Research Protection Program (HRPP) has reviewed the above referenced project and determined that it does not meet the definition of research under 45 CFR 46.102 and therefore does not require IRB approval or oversight.

If you have any questions related to this communication or the UT Southwestern HRPP, please call 214-648-3060.

**UT Southwestern Medical Center** 5323 Harry Hines Boulevard / Dallas, Texas 75390-8843 / (214)648-3060

